# Liver Metastasis Risk and Timing in Pancreatic Cancer Patients Using Electronic Health Records

**DOI:** 10.1101/2025.10.31.25339268

**Authors:** Nabasmita Talukdar, Zeshui Yu, Zheng Zeng, Xiaodan Zhang, Ying Lu, Dimitri F. Joseph, Dmitry Leshchiner, Hui Wang, Bin Chen

## Abstract

**Background:** Liver metastasis is a frequent and serious complication of pancreatic cancer, contributing to its high mortality rate. Identifying risk factors and understanding the timing of liver metastasis may improve early detection and support more effective treatment planning.

**Method:** A cohort of 12,955 incident pancreatic cancer patients was assembled from the Truveta platform. A subgroup of cases and controls that met the inclusion/exclusion criteria was analyzed using logistic regression and was reported as the primary analysis for this study. Effects on time to liver metastasis were also analyzed using univariate and multivariate Cox regression models. The primary outcome was the occurrence of liver metastasis within 1 year of diagnosis. Subjects were categorized as having baseline metastasis (≤ 30 days from pancreatic cancer diagnosis) or post-baseline metastasis (>30 days). Demographic characteristics and comorbidities were evaluated for their potential role as risk factors.

**Result:** Among 7,858 patients in the case-control cohort, 2,920 (37%) developed liver metastasis within one year, while 2,066 (70%) subjects were diagnosed with metastasis at baseline. Male sex, older age, and a history of Type 2 diabetes mellitus, depression, obesity, anemia, abdominal pain, and distant metastasis were significantly associated with a higher risk of liver metastasis. Lower odds of liver metastasis were observed among Black or African American and Hispanic or Latino subjects. In the subgroup analysis after removing baseline metastasis, surgery and radiotherapy were protective, while tumors located in the head of the pancreas showed a higher risk for metastasis in this cohort.

**Conclusions:** This study identified key clinical and demographic risk factors for liver metastasis in pancreatic cancer, emphasizing the importance of using real-world data, analyzing the timing of disease progression, and highlighting opportunities for earlier intervention and personalized care.

## Introduction

Pancreatic cancer (PC) ranks as the third leading cause of cancer-related mortality in the United States, despite being the tenth most common cancer diagnosis overall. In 2025, an estimated 67,440 new cases and 51,980 deaths are expected^1,2^. This disease poses a major public health and economic burden, with costs of USD 2.55 billion in 2016 and average expenses of USD 15,480 per patient per month^3,4^. Prognosis remains poor, with five-year survival rates ranging from 3% to 44% and median overall survival typically only 3 to 12 months, depending on the stage at diagnosis, treatment regimens, and patient characteristics^5,6,7,8,9,10^.

Diagnosis is often delayed as early-stage PC is asymptomatic or presents with nonspecific symptoms, leading most patients to seek care only after the disease has advanced^11,12^. The pancreas’s anatomical location further complicates early detection^13^. Studies have shown modest sensitivity in detecting using CT, ultrasound, or MRI, particularly for small lesions^14^. Consequently, over 50-60% of patients are diagnosed at a stage when distant metastases are already present, with the liver being the most common site of metastasis^10,11,15,16,17^. Surgery is generally not effective for metastatic disease, but even with treatment, the benefits are limited; however, some studies suggest synchronous resection may benefit carefully selected patients with PC^18^. One study reported a median time to liver metastasis (LM) of 6.0 ± 4.6 months, with 95.1% occurring within one year of pancreatectomy^19^. In metastatic PC, combination therapies such as FOLFIRINOX have been shown to extend median survival to 11.1 months, compared to 6.8 months with gemcitabine alone. The benefits of combination chemotherapies are limited by significant association with toxicity (grade 3-4 neutropenia: 46% vs. 21%)^20^.

Therefore, identifying risk factors and exploring strategies to mitigate the risk of LM are critical for improving early detection, slowing disease progression, and informing treatment decisions in patients living with PC. Molecular profiling of biopsies obtained during surgery can reveal molecular features associated with LM, but acquiring such tissue is often not feasible in daily clinical practice^21^. While clinical trials have provided valuable insights, their survival estimates may not fully generalize to real-world populations^22^.

To provide real-world evidence, we leveraged a larger Electronic Health Records (EHR) dataset collected from routine clinical practice. Our Truveta dataset includes robust demographic annotations, clinical metadata, and lab values, providing greater heterogeneity and complexity in patient characteristics. Compared to earlier studies, this allows us to describe the risk of LM more precisely, supported by our longitudinal follow-up design. Our study aims to generate a comprehensive overview of factors associated with the risk and progression of LM, as well as to evaluate differences between baseline and post-baseline LM. By utilizing individual-level data, we aim to inform future hypothesis-driven research and support the development of early identification strategies and personalized clinical decision-making.

## Method

### Data

This observational study used data compiled from the Truveta database, which includes over 120 million patients, from 30 health care organizations with an inception in 2018. Truveta provides access to continuously updated and linked EHR and claims data, including demographics, diagnoses, encounters, medications, laboratory test results, and procedures. This study used only patient records de-identified by the Health Insurance Portability and Accountability Act (HIPAA) and so did not require patient consent.

### Patient selection

We identified a total of 32,920 subjects aged 45+ years with a valid ICD-10-CM code for PC (“C25”) whose first recorded diagnosis occurred between January 1, 2019, and December 31, 2021. The index date was defined as the first recorded PC diagnosis for each subject. Subjects were required to have at least two diagnosis encounters (a valid encounter associated with a diagnosis code and a unique date) after the index date to confirm continuity of care. The diagnosis window was restricted to 2019-2021 to provide adequate follow-up periods for all subjects. The data cutoff date was November 26th, 2024.

To ensure that the identified subjects were true incident cases, we required a “quarantine” period before the first diagnosis of PC and included only subjects with at least two diagnosis encounters one year prior to the index date, one of which was an outpatient encounter recorded at least 90 days prior to the index date. Additional filters were applied, including removing subjects with records of death or secondary malignant neoplasms of the respiratory or digestive organs (ICD-10-CM “C78”) before the index date and removing subjects with missing sex information or diagnosed with endocrine PC (C25.4). The final cohort included EHR records from 12,955 subjects (full cohort). The flowchart of the study is shown in Figure 1.

**Figure 1.**
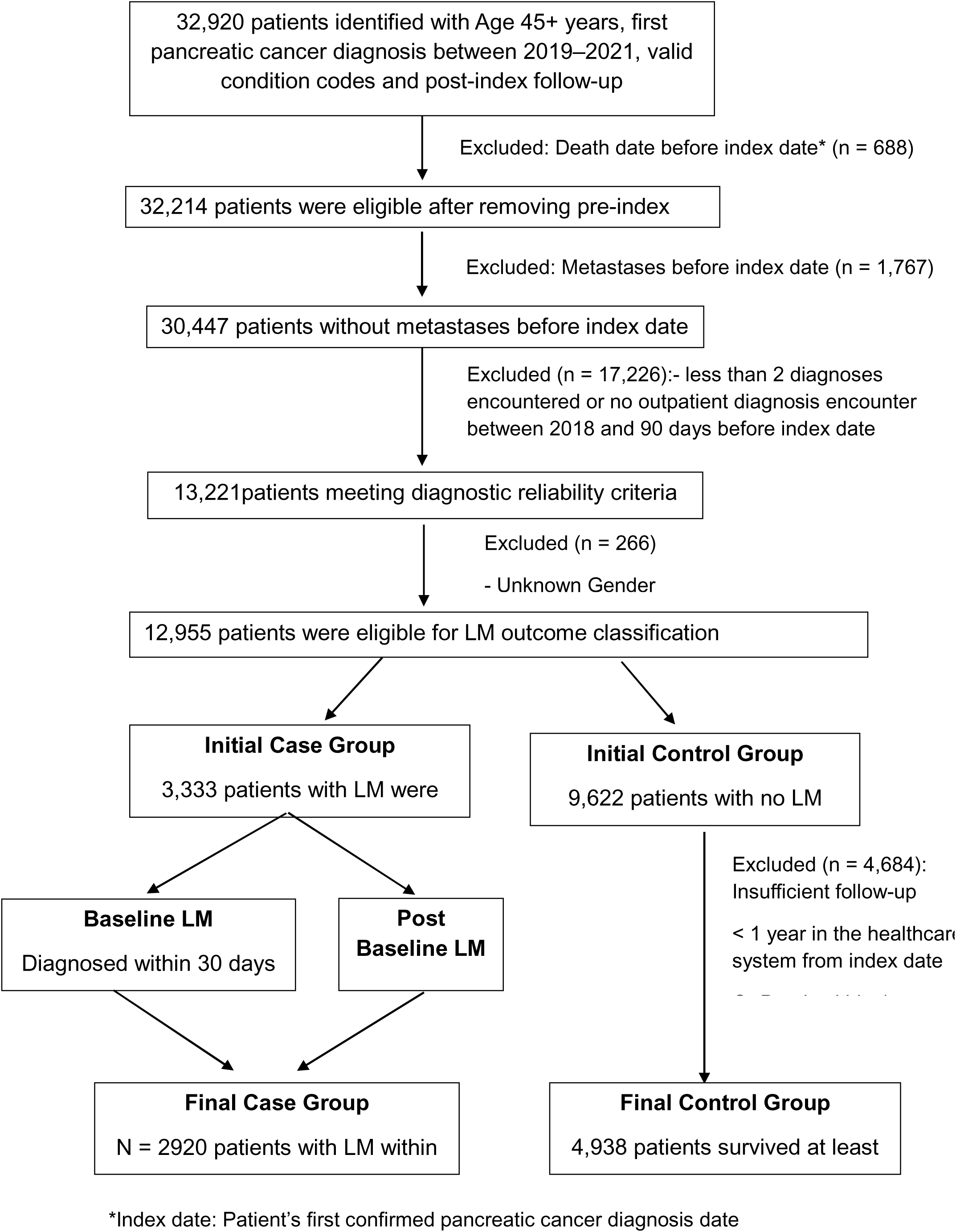
Flowchart of the study.

Pancreatic neuroendocrine tumors were excluded due to their distinct biological and clinical features compared to exocrine and ductal PC. Due to small sample sizes, some tumor locations were collapsed for analysis as follows: “head”, body and tail as “body/tail”, duct, other parts, and overlapping sites as “other”, and “unspecified”. Ungrouped counts and percentages for all primary tumor locations are presented in Figure S1.

For the primary risk factor analysis, we established a case-control cohort and limited the observation period to 1 year. The one-year observation period was chosen based on literature-reported median survival durations and time to LM progression in PC, as well as median death/LM-free survival time observed in our full cohort. The primary outcome was the development of LM (cases) within one year of the index date, identified by ICD-10-CM code “C78.7”. Subjects without documented LM who survived at least one year were classified as controls. To minimize potential misclassification of LM status, we excluded subjects who died within one year of the index date without a documented LM diagnosis. These individuals may represent cases of undiagnosed LM, as patients with advanced disease often experience rapid clinical deterioration and may not undergo further diagnostic follow-up visits. In such situations, liver metastases might not be explicitly coded in the EHR, creating a documentation gap. Excluding this group reduces the likelihood of misclassifying true LM cases as non-LM, thereby strengthening the validity of our outcome definition. This ensured a cleaner control group for comparing subjects who survived ≥1 year without LM to those who had a confirmed LM diagnosis within the first year. Additional subgroup analysis was performed using this case-control cohort by excluding LM cases within 30 days of the index date.

For the secondary analysis, we reanalyzed the full cohort after removing LM cases diagnosed within 30 days of the index date.

### Clinical variables

To account for coding delays, baseline LM was defined as any LM diagnosis occurring on or within 30 days after the index date. LM diagnoses occurring more than 30 days after the index date were classified as post-baseline LM. LM cases were divided into two groups to distinguish early from late metastasis diagnosis. Studying clinical and demographic differences between these groups could provide insights to guide treatment planning.

The follow-up time was defined as the duration from the index date to the earliest record of LM or death. For subjects without LM or death events, follow-up time was censored at the earliest of their last recorded encounter date. To account for potential under-coding of LM, we also expanded the outcome to include both LM and death for investigation in a sensitivity analysis.

Baseline characteristics (age, sex, race, ethnicity, region, comorbidities, and lab test results) were assessed during the 1-year pre-index assessment period. Comorbidities were defined using relevant diagnosis codes after excluding invalid/unknown records (Table S1). Comorbidities were selected using two different approaches: clinical relevance (e.g., known PC risk factors) and data-driven approaches (most frequent comorbidities). Distant metastases were defined as metastatic disease spreading to lymph nodes or other organs, not including the liver.

Age at diagnosis was calculated from birth date and index date, used as a continuous variable in regression models, and categorized as (45-60, 61-74, 75+) for descriptive summaries. BMI data were collected from records nearest to the index date, within a window of 1 year before to 1 year after the index date, and were retained. Missing BMI values were imputed using the multivariate imputation by chained equations (MICE) method (Figure S2). Laboratory measurements such as Direct Bilirubin, Total Bilirubin, Alkaline Phosphatase (ALP), Aspartate Aminotransferase (AST), Alanine Aminotransferase (ALT), Albumin, Total Protein, Serum Lipase, Serum Pancreatic Amylase, and Prothrombin Time (PT) were standardized to conventional units where possible. Values with missing or incompatible (such as second vs. IU/L) units or exceeding predefined physiologic plausibility thresholds were excluded.

### Treatment variables

To evaluate risk factors for post-index treatments (chemotherapy, radiotherapy, surgery), analysis was limited to subjects without baseline LM. Chemotherapy exposure was defined by at least two records from prescription, dispensation, or administration records from 30 days before the index date to the end of follow-up. Surgery and radiotherapy were identified using relevant procedure codes within the same period. The 30-day pre-index window accounted for coding lags and documentation gaps.

### Statistical Analysis

We summarized demographic and clinical characteristics using counts and percentages for categorical variables, and medians with interquartile ranges (IQR) for continuous variables. The primary analysis examined the risk of LM within one year of PC diagnosis in the case-control cohort. For subjects without LM at baseline, subgroup analyses were performed to assess the risk of developing incident LM. Odds ratios (OR) were estimated using univariate logistic regression for all variables, followed by multivariate logistic regression with stepwise selection in both directions to identify potential risk factors in both the primary and subgroup analysis case-control cohorts.

For the secondary analysis, we used the full cohort after excluding baseline LM cases. LM was defined as the event of interest, while death, loss to follow-up, or end of observation were treated as censoring. Time-to-event outcomes were analyzed using multivariate Cox proportional hazards models with stepwise selection in both directions to identify significant predictors of LM risk over time. As a sensitivity analysis, we applied a composite endpoint of LM or death to account for patients who died without a documented LM diagnosis, again using multivariate Cox regression with stepwise selection in both directions.

Among LM cases, the Kruskal-Wallis test assessed differences in median time to metastasis across clinical and demographic subgroups. Chi-square tests compared baseline and post-baseline LM characteristics, while Wilcoxon rank-sum tests evaluated lab test values between LM cases and controls. All tests were two-sided, with significance set at p < 0.05. The median follow-up time for the full cohort was estimated using the reverse Kaplan-Meier method.

## Results

In the full cohort of 12,955 subjects, a total of 3,333 (25.7%) subjects with LM were identified, of which only 1,267 (38%) were post-baseline LM. Among these post-baseline LM subjects, the median time to LM was 0.6 years (IQR: 0.3, 1.2). A total of 8,618 (66%) subjects died, of which 7,768 (90%) died within the first year of PC diagnosis. The median follow-up time in the full cohort is 3.55 years (95% Confidence Interval (CI): 3.51, 3.61).

Of the 7,858 subjects in the case-control sub-cohorts, 2,920 (37%) developed LM within 1 year of the index date. The median age at diagnosis was 71 years, with 50% of subjects falling within the 61-74 age group. Only a small fraction of subjects (223, 3%) identified as Hispanic or Latino. The majority (76%) were white, 7.6% were Black or African American, and the remaining 12% were recorded as unknown. The sex distribution was balanced, with females comprising 52% of the population. Common comorbidities included abdominal pain (59%), hyperlipidemia (72%), type 2 diabetes mellitus (T2DM) (42%), anemia (39%), and hypertension (77%). A complete descriptive table with univariate results is provided in Table 1. Figure 2 presents a forest plot of odds ratios from the multivariate logistic regression models for LM, stratified by baseline LM status.

**Figure 2:**
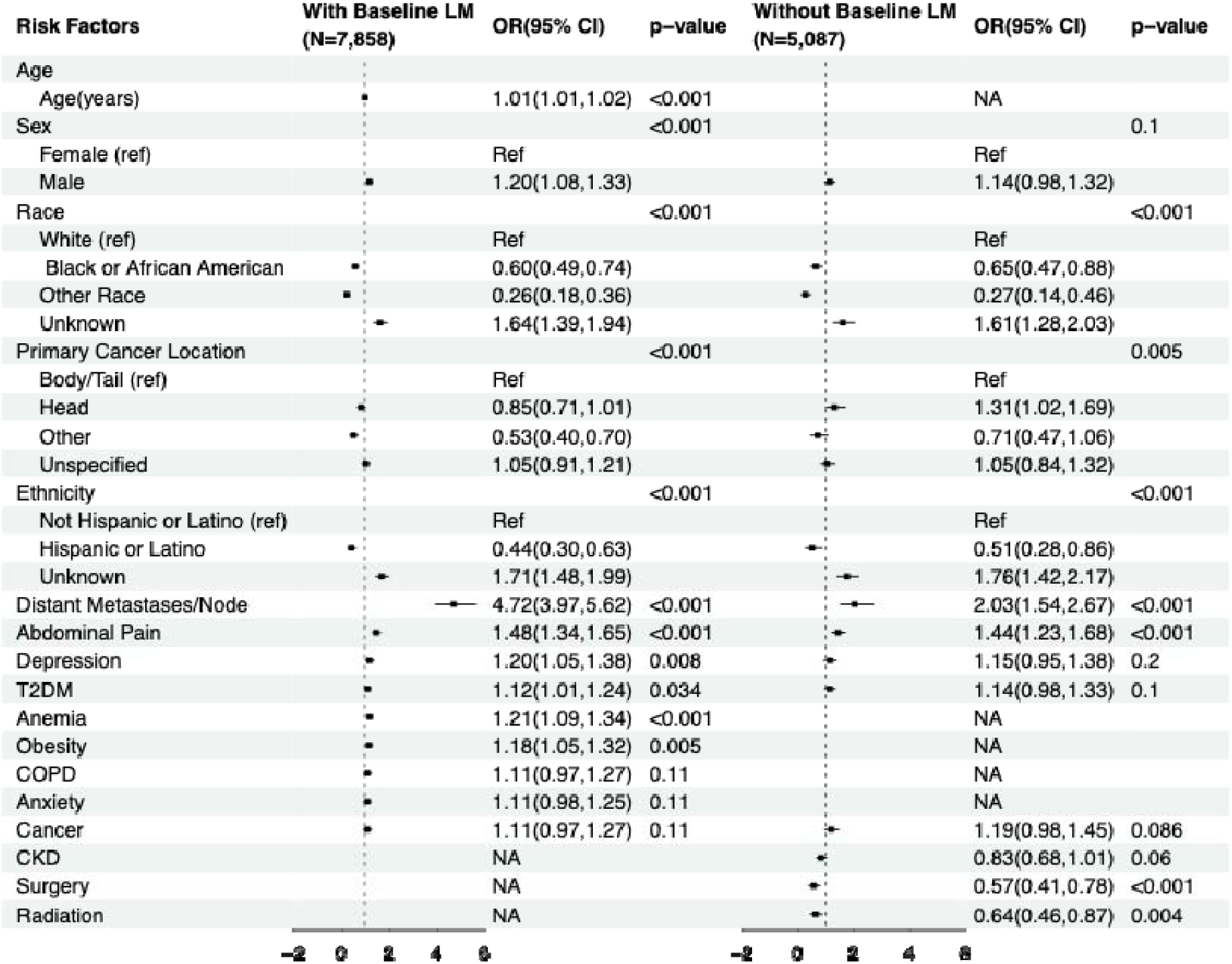
Forest Plot of Multivariate Logistic Regression Analysis for LM Risk Factors for the Case-Control Cohort.

**Table 1:** Patient Characteristics and Univariate Logistic Regression Analysis for Risk of Liver Metastasis in Case-Control Cohort.

| Characteristic | Descriptive Statistics |  |  | Univariate Logistic Regression |  |
| --- | --- | --- | --- | --- | --- |
|  | Overall<br>N = 7,858 | Control<br>N = 4,938 | Cases<br>N = 2,920 | OR | p-value |
| **Age (years), Median (Q1, Q3)** | 71 (64, 78) | 70 (63, 77) | 71 (64, 78) | 1 | <0.001 |
| **Age Group, n (%)** |  |  |  | NA | NA |
| 45-60 | 1,159 (15%) | 783 (16%) | 376 (13%) |  |  |
| 61-74 | 3,873 (49%) | 2,459 (50%) | 1,414 (48%) |  |  |
| 75+ | 2,826 (36%) | 1,696 (34%) | 1,130 (39%) |  |  |
| **Ethnicity, n (%)** |  |  |  |  | <0.001 |
| Not Hispanic or Latino | 6,403 (81%) | 4,175 (85%) | 2,228 (76%) | Ref |  |
| Hispanic or Latino | 250 (3.2%) | 211 (4.3%) | 39 (1.3%) | 0.83 |  |
| Unknown | 1,205 (15%) | 552 (11%) | 653 (22%) | 1.21 |  |
| **Race, n (%)** |  |  |  |  | <0.001 |
| White | 5,951 (76%) | 3,745 (76%) | 2,206 (76%) | Ref |  |
| Black or African American | 613 (7.8%) | 456 (9.2%) | 157 (5.4%) | 0.89 |  |
| Other Race | 341 (4.3%) | 301 (6.1%) | 40 (1.4%) | 0.78 |  |
| Unknown | 953 (12%) | 436 (8.8%) | 517 (18%) | 1.19 |  |
| **Sex, n (%)** |  |  |  |  | 0.004 |
| Female | 4,059 (52%) | 2,613 (53%) | 1,446 (50%) | Ref |  |
| Male | 3,799 (48%) | 2,325 (47%) | 1,474 (50%) | 1.03 |  |
| **Region, n (%)** |  |  |  | NA | NA |
| Midwest | 682 (8.7%) | 575 (12%) | 107 (3.7%) |  |  |
| Northeast | 279 (3.6%) | 232 (4.7%) | 47 (1.6%) |  |  |
| South | 941 (12%) | 824 (17%) | 117 (4.0%) |  |  |
| Unknown | 5,289 (67%) | 2,773 (56%) | 2,516 (86%) |  |  |
| West | 667 (8.5%) | 534 (11%) | 133 (4.6%) |  |  |
| **Distant Metastases/Node, n (%)** |  |  |  |  | <0.001 |
| No | 7,059 (90%) | 4,708 (95%) | 2,351 (81%) | Ref |  |
| Yes | 799 (10%) | 230 (4.7%) | 569 (19%) | 1.46 |  |
| **Primary Cancer Location, n (%)** |  |  |  |  | <0.001 |
| Body/Tail | 1,113 (14%) | 689 (14%) | 424 (15%) | Ref |  |
| Head | 1,492 (19%) | 995 (20%) | 497 (17%) | 0.95 |  |
| Other | 422 (5.4%) | 315 (6.4%) | 107 (3.7%) | 0.88 |  |
| Unspecified | 4,831 (61%) | 2,939 (60%) | 1,892 (65%) | 1.01 |  |
| **BMI, Median (Q1, Q3)** | 26.6 (23.2, 30.7) | 26.6 (23.4, 30.7) | 26.5 (23.0, 30.8) | 1 | 0.5 |
| **BMI Group, n(%)** |  |  |  | NA | NA |
| Normal | 2,682 (34%) | 1,633 (33%) | 1,049 (36%) |  |  |
| □□□□ Underweight | 273 (3.5%) | 176 (3.6%) | 97 (3.3%) | □ | □ |
| □□□□ Overweight | 2,674 (34%) | 1,747 (35%) | 927 (32%) | □ | □ |
| □□□□ Obese (Class I) | 1,339 (17%) | 819 (17%) | 520 (18%) | □ | □ |
| □□□□ Obese (Class II) | 538 (6.8%) | 342 (6.9%) | 196 (6.7%) | □ | □ |
| □□□□ Obese (Class III) | 352 (4.5%) | 221 (4.5%) | 131 (4.5%) | □ | □ |
| **CKD, n (%)** | 1,629 (21%) | 952 (19%) | 677 (23%) | 1.06 | <0.001 |
| **T2DM, n (%)** | 3,305 (42%) | 1,994 (40%) | 1,311 (45%) | 1.04 | <0.001 |
| **Hepatitis, n (%)** | 222 (2.8%) | 133 (2.7%) | 89 (3.0%) | 1.03 | 0.4 |
| **Hypertension, n (%)** | 6,020 (77%) | 3,703 (75%) | 2,317 (79%) | 1.06 | <0.001 |
| **CLD, n (%)** | 339 (4.3%) | 192 (3.9%) | 147 (5.0%) | 1.07 | 0.016 |
| **Hyperlipidemia, n (%)** | 5,662 (72%) | 3,505 (71%) | 2,157 (74%) | 1.03 | 0.006 |
| **OSA, n (%)** | 1,456 (19%) | 897 (18%) | 559 (19%) | 1.02 | 0.3 |
| **COPD, n (%)** | 1,301 (17%) | 753 (15%) | 548 (19%) | 1.06 | <0.001 |
| **Anxiety, n (%)** | 2,086 (27%) | 1,248 (25%) | 838 (29%) | 1.04 | <0.001 |
| **IHD, n (%)** | 2,032 (26%) | 1,200 (24%) | 832 (28%) | 1.05 | <0.001 |
| **Depression, n (%)** | 1,556 (20%) | 897 (18%) | 659 (23%) | 1.07 | <0.001 |
| **Obesity, n (%)** | 2,169 (28%) | 1,295 (26%) | 874 (30%) | 1.04 | <0.001 |
| **Other Cancer, n (%)** | 1,374 (17%) | 812 (16%) | 562 (19%) | 1.05 | 0.002 |
| **Abdominal Pain, n (%)** | 4,614 (59%) | 2,723 (55%) | 1,891 (65%) | 1.1 | <0.001 |
| **Dyspnea, n (%)** | 2,253 (29%) | 1,349 (27%) | 904 (31%) | 1.04 | <0.001 |
| **Anemia, n (%)** | 3,080 (39%) | 1,801 (36%) | 1,279 (44%) | 1.07 | <0.001 |

Abbreviations: CI = Confidence Interval, HR = Hazard Ratio, NA□ = Not Applicable, IHD = Ischemic Heart Disease, OSA = Obstructive sleep apnea, COPD = Chronic obstructive pulmonary disease, FHOMND = Family History of Malignant Neoplasm of Digestive System, T2DM = Type 2 Diabetes Mellitus, BMI = Body Mass Index, CKD = Chronic Kidney Disease, CLD = Chronic Liver Disease, Obese (Class I): 30 ≤ BMI < 35, Obese (Class II): 35 ≤ BMI < 40, Obese (Class III): 40 ≥ BMI

In the multivariate logistic regression model for primary analysis, several factors were significantly associated with increased odds of LM. These included older age (OR = 1.01, 95% CI = 1.01-1.02, p < 0.001), male sex (OR = 1.20, 95% CI = 1.08-1.33, p < 0.001), T2DM(OR = 1.12, 95% CI = 1.01-1.24, p = 0.034), abdominal pain (OR = 1.48, 95% CI 1.34-1.65, p < 0.001), depression (OR = 1.20, 95% CI = 1.05-1.38, p = 0.008), anemia (OR = 1.21, 95% CI = 1.09-1.34, p < 0.001), obesity (OR = 1.18, 95% CI = 1.05-1.32, p = 0.005), and distant metastases or lymph node involvement (OR = 4.72, 95% CI = 3.97-5.62, p < 0.001). Lower odds of LM were observed among Black (OR = 0.60, 95% CI = 0.49-0.74) and Hispanic or Latino (OR = 0.44, 95% CI = 0.30-0.63) subjects, compared to White and non-Hispanic counterparts (p < 0.001). Tumors in the pancreatic head showed a trend toward reduced odds compared to body/tail tumors (OR = 0.85, 95% CI = 0.71-1.01), though the significance was borderline. Overall, tumor location was significantly associated with outcome (global p < 0.001).

In the subgroup analysis after removing LM at baseline, higher odds of LM were significantly observed with tumors in the pancreatic head (OR = 1.31, 95% CI = 1.02-1.69), distant metastases or lymph node involvement (OR = 2.03, 95% CI = 1.54-2.67), and abdominal pain (OR =1.44, 95% CI = 1.23-1.68). On the other hand, Hispanic or Latino ethnicity (OR = 0.51, 95% CI = 0.28-0.86) and Black race (OR =0.65, 95% CI = 0.47-0.88) compared to White Race were associated with lower odds of LM. Surgery (OR =0.57, 95% CI = 0.4-0.78, p < 0.001) and radiation (OR = 0.64, 95% CI = 0.46-0.87, p = 0.004) were also significantly associated with lower odds of post-baseline LM.

In the time-to-event Cox regression model (N = 10,889), we observed consistent trends with the earlier logistic regression analysis. Additional covariates included anemia (HR = 0.88, 95% CI = 0.78-0.99) and other cancer (HR = 1.17, 95% CI = 1.01-1.35), as shown in Table S2. In the sensitivity analysis, where a composite outcome of LM and death was used, further variables entered the model, including T2DM (HR = 1.18, 95% CI =1.13-1.24), obesity (HR = 1.19, 95% CI = 1.11-1.26), chronic obstructive pulmonary disease (COPD) (HR = 1.16, 95% CI: 1.09-1.23), Obstructive sleep apnea(OSA) (HR = 0.93, 95% CI = 0.87-1.00), CLD (HR = 0.26, 95% CI = 0.13-0.41), hyperlipidemia (HR = 0.92, 95% CI: = 0.87-0.95), and radiation (HR = 0.87, 95% CI = 0.71-1.08). Kaplan-Meier survival curves illustrating time to LM and the composite outcome (LM or death) are presented in Figure S3.

The time to LM was summarized descriptively by baseline variables among 1,267 post-baseline LM subjects Table S3. Median time to LM varied across demographics, treatment exposures, and comorbidities. Subjects who received surgery (1.10 vs. 0.56 years), radiation (1.13 vs. 0.55 years), or chemotherapy (0.90 vs. 0.43 years) had longer times to metastasis compared to those who didn’t.

Chi-square tests comparing baseline vs. post-baseline LM groups showed distinct clinical differences (Table S4). Baseline LM cases were aged 75+ (40.9% vs. 31.3%), had more body/tail tumors (15.2% vs. 13%), and more distant/lymph node metastases (23.7% vs. 9.7%), whereas post-baseline LM had more patients aged 45-60 (17.2% vs. 11.2%) and more head tumors (27.2% vs. 13.6%). Baseline LM cases had higher rates of chronic kidney disease (CKD) (25.5% vs. 17.3%), chronic liver disease (CLD) (5.5% vs. 3.3%), hyperlipidemia (75.4% vs. 69.2%), COPD (19.4% vs. 15.5%), ischemic heart disease (IHD) (29.6% vs. 23.6%), and anemia (47.0% vs. 34.4%).

Exploratory laboratory data analysis results are shown in Table S5. It showed that LM cases had significantly higher median values of direct bilirubin, total bilirubin, ALP, AST, ALT, and PT compared to the control group. In contrast, albumin and total protein were lower in LM cases compared to controls.

## Discussion

This study is among the first EHR-based investigations to evaluate the timing and risk factors of LM in patients with PC. We identified 32,920 subjects aged 45 years or older with PC. After applying data quality filters to address data incompleteness, we assembled a cohort of 12,955 subjects with incident PC for analysis. Within this cohort, 3,333 subjects were identified with LM, and 8,618 subjects died at any time during the observation period.

Our findings suggest that the development of LM in PC is complex and multifactorial, influenced by demographic, biological, and treatment-related factors. Older age, male sex, White race, distant metastases or lymph node involvement, T2DM, depression, obesity, anemia, and abdominal pain were significantly associated with higher LM risk. Cancer, anxiety, and COPD showed higher odds of LM, but were not statistically significant. After excluding baseline LM to assess treatment exposures, the multivariate model showed similar trends, with surgery and radiation demonstrating protective effects. Depression and T2DM remained in the model but did not retain statistical significance, while anemia and obesity were not present in the model. Previous studies have reported similar trends regarding age, sex, and treatment; however, our findings on race differ from prior work^23,24^. In addition, we were unable to identify any previous studies that reported results related to comorbidities, highlighting that our study provides the first evidence of its kind.

Tumor location also influenced LM risk, though associations varied by cohort stratification. In the full cohort, head tumors had a lower LM risk compared to body/tail tumors. This aligns with prior studies showing that body/tail tumors have higher odds of LM, despite the pancreatic head being the most common tumor site, mostly due to late diagnosis of body/tail tumors^8,23,24,25,26^. However, after excluding baseline LM, head tumors showed a higher subsequent risk, likely due to body/tail tumors contributing more baseline LM cases being removed. This finding underscores the importance of distinguishing baseline from incident metastases and suggests that patients without baseline LM and a tumor at the head of the pancreas may represent a subgroup at particularly elevated risk of LM.

Time-to-event analysis on the full cohort showed a trend consistent with the primary findings, with additional covariates included in the sensitivity model to evaluate the composite outcome of LM or death. For instance, anemia, which showed higher odds in earlier models, appeared protective in the Cox model for time to LM, though with a marginal CI. Similarly, OSA and hyperlipidemia showed a lower hazard ratio, likely due to residual confounding, as lipid-lowering medications may be effective in slowing cancer metastasis^27,28^.

The consistent findings that surgery and radiation therapy are associated with a reduced risk of LM across all models strengthen the robustness of our results and highlight their therapeutic significance in early-stage management^29^. Individuals aged 75+ had higher baseline LM rates, suggesting delayed diagnosis, aggressive disease, or underuse of screening in older adults. Baseline LM cases also had a greater comorbidity burden (CKD, hypertension, CLD, hyperlipidemia, IHD, dyspnea, anemia, COPD). The observed pattern may reflect the challenges in achieving accurate and timely PC diagnosis, partially resulting from the variability and overlap of clinical signs and symptoms or prioritized management in patients with multiple comorbidities, which can complicate recognition of the underlying malignancy. Prior studies have shown that multimorbidity can delay cancer diagnosis through reduced diagnostic intensity or misattribution of symptoms to chronic illness^30,31^.

Exploratory analyses on laboratory measurements showed that LM cases had elevated liver function markers (direct bilirubin, ALP, AST, ALT, PT) and lower albumin and total protein levels. Consistent with prior research, these biomarkers may signal early physiological changes related to liver involvement and may prove valuable in evaluating disease severity ^23^. However, due to incomplete lab data and potential selection bias, laboratory test measurements were excluded from the primary models. Nonetheless, further research is warranted to investigate the potential role of these laboratory markers in LM risk stratification.

Previous studies have examined prognostic factors, incidence, and survival of LM; assessed various machine learning models to predict LM risk; or explored associations between laboratory markers and LM in advanced PC patients^17,23,24,32^. However, most of these studies focused on overall survival, lacked detailed comorbidity and treatment data, or analyzed LM at a single time point, limiting assessment of temporal and clinical factors driving LM development. We applied stricter inclusion criteria and complementary statistical approaches to obtain more comprehensive evidence on the development of LM: logistic regression on a clean case-control cohort and time-to-event analysis on the full cohort, including a sensitivity analysis with a composite outcome of LM or death to mitigate under-capture bias. Incorporating recent clinical data from routine medical practice offers multiple benefits, including capturing diverse real-world patient groups, reflecting modern diagnostic methods and treatment variability, enabling long-term follow-up, supporting hypothesis development, and enhancing the generalizability of results.

Our result should be interpreted in the context of a few limitations. The observational nature of this study limits the ability to draw any causal conclusion, and the potential for residual confounding cannot be excluded. We are lacking cancer staging information (e.g., TNM or AJCC stage), which could have allowed the determination of the baseline severity of PC. Although we used surgery and distant metastases as a proxy for early-stage disease, misclassification is possible. Socioeconomic factors, which are known to influence access to care, timing of diagnosis, and follow-up intensity, were not fully captured in our analysis and warrant further investigation. Additionally, the identification of PC and LM was based on ICD codes rather than imaging or pathology confirmation, although prior research supports the validity of ICD coding for disease identification^33^.

## Conclusion

In conclusion, this study highlights real-world patterns in the incidence and timing of LM among patients living with PC. By distinguishing baseline from post-baseline metastasis and evaluating clinical factors, our findings improve risk stratification and demonstrate the utility of EHR data for metastatic cancer surveillance and care planning. Although EHR data may under-report events due to documentation gaps, we successfully identified key LM risk factors. This underscores how EHR-based research, when combined with rigorous data curation and thoughtful analytics, can yield clinically meaningful insights.

## Acknowledgements

The authors would like to acknowledge and thank Angela Pei from the University of California, Santa Barbara, for her assistance with creating the forest plot in this manuscript.

The funders had no role in the design and conduct of the study; collection, management, analysis, and interpretation of the data; preparation, review, or approval of the manuscript; and decision to submit the manuscript for publication.

During the preparation of this work, the author(s) used ChatGPT (OpenAI, GPT-4) to assist with language editing and improvement of readability. After using this tool, the author(s) reviewed and edited the content as needed and take(s) full responsibility for the content of the published article.

## Funding/Support

The research is supported by the NIH R01GM134307, R01GM145700 and the MSU SPG grant.

## Conflicts of interest

The authors have no conflicts to disclose.

## Data Availability

Data cannot be shared for ethical/privacy reasons. As a result of contractual agreements with TRUVETA, we are unable to export individual patient data. It is possible for researchers to gain access to individual patient data, but they must enter into an agreement with TRUVETA and pay a fee in order to do so. The data used in this study is available to all Truveta subscribers and may be accessed at studio.truveta.com.

## Notes

### Competing Interest Statement

The authors have declared no competing interest.

### Summary of Updates

The manuscript currently doesn't have any main figures.

