## SupplementaryFile for "Liver Metastasis Risk and Timing in Pancreatic Cancer Patients Using Electronic Health Records"

**Supplemental Content**

**Table S1:** Comorbidities with ICD codes

**Table S2:** Multivariate Cox Regression Analysis on the whole cohort, along with sensitivity analysis after removing baseline LM

**Table S3:** Summary of Time to Liver Metastasis Among Post-Baseline Cases

**Table S4:** Comparative Characteristics for Baseline vs. Post-Baseline Liver Metastasis Along with Patient Characteristics for All Controls

**Table S5:** Distribution of Lab Function Test for Liver Metastasis (Cases) and Subjects Free of Liver Metastases (Control)

**Figure S1**: Frequency of Primary Pancreatic Cancer Location

**Figure S2:** Distribution of observed and imputed BMI values.

**Figure S3:** Kaplan-Meier survival curves illustrating time to LM and the composite outcome (LM or death)

**Table 1: Comorbidities with ICD codes**

| **Comorbidities** | ICD10 CM Codes |
| --- | --- |
| **Chronic Kidney Disease** | "E13.22", "E11.22", "E08.22", "E09.22", "N18", "I12", "I13" |
| **T2DM** | "E11" |
| **Hepatitis (B & C)** | "B16.0", "B16.1", "B16.2", "B16.9", "B17.0", "B18.0", "B18.1", "B19.10", "B19.11", "B17.1", "B17.10", "B17.11", "B18.2", "B19.2", "B19.20", "B19.21" |
| **Chronic Liver Disease** | "K70.0", "K70.2", "K70.30", "K70.31", "K70.40", "K70.41", "K70.9", "K71.0", "K71.10", "K71.11", "K71.3", "K71.4", "K71.50", "K71.51", "K71.7", "K72.10", "K72.11", "K73.0", "K73.1", "K73.2", "K73.8", "K73.9", "K74.0", "K74.00", "K74.01", "K74.02", "K74.1", "K74.2", "K74.3", "K74.4", "K74.5", "K74.60", "K74.69", "K76.1", "K76.2", "K76.3", "K76.5", "K76.6", "K76.7" |
| **Hyperlipidemia** | "E78.0", "E78.00", "E78.01", "E78.1", "E78.2", "E78.3", "E78.4", "E78.41", "E78.49", "E78.5" |
| **Hypertension** | "I10", "I11", "I12", "I13", "I15", "I16" |
| **Anxiety** | "F41.0", "F41.1", "F41.3", "F41.8", "F41.9" |
| **Depression** | "F01.51", "F32.0", "F32.1", "F32.2", "F32.3", "F32.4", "F32.5", "F32.89", "F32.9", "F33", "F34.1", "F34.81", "F34.89", "F43.21", "F43.23", "F53.0", "F53.1" |
| **Ischemic Heart Ischemic** | "I23.0", "I23.1", "I23.2", "I23.3", "I23.4", "I23.5", "I23.6", "I23.8", "I24.0", "I24.8", "I24.9", "I25" |
| **Obesity** | "E66.01", "E66.09", "E66.1", "E66.2", "E66.3", "E66.8", "E66.9" |
| **Abdominal Pain** | "R10.9", "R10.30", "R10.10", "R10.84", "R10.11", "R10.31", "R10.32", "R10.8" |
| **Dyspnea** | "R06.00", "R06.0", "R06.09", "J95.87", "R06.02" |
| **Anemia** | "D50.0", "D50.1", "D50.8", "D50.9", "D55.0", "D55.1", "D55.21", "D55.29", "D55.3", "D55.8", "D55.9", "D58.0", "D58.8", "D58.9", "D59.0", "D59.10", "D59.11", "D59.12", "D59.13", "D59.19", "D59.2", "D59.3", "D59.4", "D59.5", "D59.6", "D59.8", "D59.9", "D63.0", "D63.1", "D63.8", "O99.011", "O99.012", "O99.013", "O99.019", "O99.02", "D51.0", "D51.3", "D51.8", "D51.9", "D53.9", "D62", "D64.81", "D64.89", "D64.9", "O90.81", "O99.01" |
| **COPD** | "J41", "J42", "J43", "J44" |
| **Obstructive Sleep Apnea (OSA)** | "G47.3", "G47.30", "G47.33", "G47.39" |
| **Cancer (Breast, Lung, Esophageal, Stomach, Colorectal)** | "C15", "C16","C18","C19","C20", "C21", "C22”, “C33","C34", "C50" |

**Table 2: Multivariate Cox Regression Analysis on the whole cohort, along with sensitivity analysis after removing baseline LM**

|  | **Multivariate Cox Regression^1^, N = 10,889** | | | **Sensitivity Multivariate Cox Regression^2^, N = 10,889** | | |
| --- | --- | --- | --- | --- | --- | --- |
| **Characteristic** | **HR** | **95% CI** | **p-value** | **HR** | **95% CI** | **p-value** |
| **Age (years)** | 1.00 | 0.99, 1.00 | 0.018 | 1.03 | 1.02, 1.03 | <0.001 |
| **Sex** |  |  | 0.016 |  |  | 0.013 |
| Female | — | — |  | — | — |  |
| Male | 1.15 | 1.03, 1.28 |  | 1.06 | 1.01, 1.12 |  |
| **Ethnicity** |  |  | <0.001 |  |  | <0.001 |
| Not Hispanic or Latino | — | — |  | — | — |  |
| Hispanic or Latino | 0.53 | 0.33, 0.83 |  | 0.13 | 0.08, 0.20 |  |
| Unknown | 1.64 | 1.41, 1.91 |  | 1.62 | 1.52, 1.73 |  |
| **Race** |  |  | <0.001 |  |  | <0.001 |
| White | — | — |  | — | — |  |
| *Black or African American* | 0.68 | 0.54, 0.87 |  | 0.61 | 0.55, 0.68 |  |
| Other Race | 0.36 | 0.23, 0.56 |  | 0.09 | 0.06, 0.13 |  |
| Unknown | 1.26 | 1.07, 1.50 |  | 1.49 | 1.39, 1.59 |  |
| **Distant Metastases/Node** | 1.59 | 1.32, 1.93 | <0.001 | 1.69 | 1.56, 1.84 | <0.001 |
| **BMI** |  |  |  | 0.99 | 0.98, 0.99 | <0.001 |
| **Primary Cancer Location** |  |  | 0.002 |  |  | <0.001 |
| Body/Tail | — | — |  | — | — |  |
| Head | 1.26 | 1.04, 1.52 |  | 1.16 | 1.07, 1.26 |  |
| Other | 0.83 | 0.62, 1.12 |  | 0.76 | 0.67, 0.86 |  |
| Unspecified | 1.02 | 0.86, 1.22 |  | 1.1 | 1.02, 1.18 |  |
| **Anemia** | 0.88 | 0.78, 0.99 | 0.028 | 1.06 | 1.01, 1.11 | 0.026 |
| **Cancer** | 1.17 | 1.01, 1.35 | 0.037 |  |  |  |
| **Abdominal Pain** | 1.29 | 1.14, 1.44 | <0.001 | 1.12 | 1.07, 1.18 | <0.001 |
| **T2DM** |  |  |  | 1.18 | 1.13, 1.24 | <0.001 |
| **COPD** |  |  |  | 1.16 | 1.09, 1.23 | <0.001 |
| **Obesity** |  |  |  | 1.19 | 1.11, 1.26 | <0.001 |
| **CLD** |  |  |  | 1.26 | 1.13, 1.41 | <0.001 |
| **Hyperlipidemia** |  |  |  | 0.92 | 0.87, 0.98 | 0.005 |
| **OSA** |  |  |  | 0.93 | 0.87, 1.00 | 0.035 |
| **Dyspnea** |  |  |  | 1.05 | 1.00, 1.11 | 0.058 |
| **Depression** |  |  |  | 1.05 | 0.98, 1.11 | 0.2 |
| **Radiation** | 0.87 | 0.71, 1.06 | 0.2 | 0.83 | 0.75, 0.91 | <0.001 |
| **Surgery** | 0.77 | 0.63, 0.95 | 0.012 | 0.59 | 0.52, 0.66 | <0.001 |
| 1: Liver metastases as event of interest and Death is censored  2: Both Liver metastases and death are considered as event | | | | | | |

**Table 3:** **Summary of Time to Liver Metastasis Among Post-Baseline Cases**

|  | **Median, N = 1267** |
| --- | --- |
| **Characteristic** | **Years (IQR)***^1^* |
| **Age Group, n (%)** |  |
| *Age 45-60* | 0.64 (0.28, 1.27) |
| *Age 61-74* | 0.67 (0.28, 1.25) |
| *Age >= 75* | 0.52 (0.24, 1.12) |
| **Ethnicity, n (%)** |  |
| *Not Hispanic or Latino* | 0.60 (0.25, 1.20) |
| *Hispanic or Latino* | 0.69 (0.54, 1.46) |
| *Unknown* | 0.64 (0.28, 1.26) |
| **Race, n (%)** |  |
| *White* | 0.60 (0.25, 1.21) |
| *Black or African American* | 0.58 (0.28, 1.32) |
| *Other Race* | 0.76 (0.48, 1.87) |
| *Unknown* | 0.61 (0.30, 1.14) |
| **Sex, n (%)** |  |
| *Female* | 0.56 (0.27, 1.19) |
| *Male* | 0.65 (0.24, 1.25) |
| **Region, n (%)** |  |
| *Midwest* | 0.58 (0.20, 1.18) |
| *Northeast* | 0.60 (0.35, 1.74) |
| *South* | 0.77 (0.34, 1.62) |
| *Unknown* | 0.59 (0.26, 1.19) |
| *West* | 0.68 (0.28, 1.30) |
| **Distant Metastases/Node, n (%)** |  |
| *N* | 0.61 (0.26, 1.20) |
| *Y* | 0.57 (0.25, 1.30) |
| **Primary Cancer Location, n (%)** |  |
| *Body/Tail* | 0.53 (0.23, 1.19) |
| *Head* | 0.68 (0.28, 1.27) |
| *Other* | 0.74 (0.36, 1.46) |
| *Unspecified* | 0.56 (0.24, 1.16) |
| **BMI Group, n (%)** |  |
| *Normal* | 0.56 (0.23, 1.16) |
| *Underweight* | 0.35 (0.15, 1.08) |
| *Overweight* | 0.60 (0.27, 1.18) |
| *Obese (Class I)* | 0.69 (0.31, 1.35) |
| *Obese (Class II)* | 0.60 (0.26, 1.22) |
| *Obese (Class III)* | 0.73 (0.37, 1.46) |
| **Chronic Kidney, n (%)** |  |
| *0* | 0.62 (0.27, 1.24) |
| *1* | 0.52 (0.23, 1.15) |
| **T2DM, n (%)** |  |
| *0* | 0.63 (0.25, 1.25) |
| *1* | 0.57 (0.28, 1.16) |
| **Hepatitis, n (%)** |  |
| *0* | 0.62 (0.26, 1.22) |
| *1* | 0.35 (0.18, 0.78) |
| **Hypertension, n (%)** |  |
| *0* | 0.66 (0.24, 1.30) |
| *1* | 0.59 (0.27, 1.18) |
| **Chronic Liver, n (%)** |  |
| *0* | 0.62 (0.26, 1.22) |
| *1* | 0.41 (0.24, 0.74) |
| **Hyperlipidemia, n (%)** |  |
| *0* | 0.59 (0.24, 1.28) |
| *1* | 0.62 (0.27, 1.17) |
| **OSA, n (%)** |  |
| *0* | 0.59 (0.24, 1.20) |
| *1* | 0.65 (0.32, 1.24) |
| **COPD, n (%)** |  |
| *0* | 0.64 (0.27, 1.25) |
| *1* | 0.43 (0.19, 1.00) |
| **Anxiety, n (%)** |  |
| *0* | 0.58 (0.26, 1.18) |
| *1* | 0.68 (0.26, 1.29) |
| **IHD, n (%)** |  |
| *0* | 0.64 (0.27, 1.26) |
| *1* | 0.50 (0.24, 1.10) |
| **Depression, n (%)** |  |
| *0* | 0.61 (0.24, 1.20) |
| *1* | 0.60 (0.29, 1.22) |
| **Obesity, n (%)** |  |
| *0* | 0.60 (0.25, 1.20) |
| *1* | 0.62 (0.28, 1.25) |
| **Cancer, n (%)** |  |
| *0* | 0.62 (0.25, 1.20) |
| *1* | 0.57 (0.29, 1.22) |
| **Abdominal Pain, n (%)** |  |
| *0* | 0.60 (0.25, 1.29) |
| *1* | 0.61 (0.27, 1.17) |
| **Dyspnea, n (%)** |  |
| *0* | 0.62 (0.27, 1.23) |
| *1* | 0.59 (0.24, 1.17) |
| **Anemia, n (%)** |  |
| *0* | 0.64 (0.27, 1.26) |
| *1* | 0.55 (0.24, 1.14) |
| **Radiation, n (%)** |  |
| *0* | 0.55 (0.24, 1.17) |
| *1* | 1.13 (0.73, 1.48) |
| **Surgery, n (%)** |  |
| *0* | 0.56 (0.25, 1.17) |
| *1* | 1.10 (0.56, 1.58) |
| **Chemotherapy, n (%)** |  |
| *0* | 0.43 (0.19, 0.98) |
| *1* | 0.90 (0.44, 1.45) |
| *^1^*Time to Metastases: Median (Q1, Q3)  *^2^*Kruskal-Wallis rank sum test; Wilcoxon rank sum test  IHD = Ischemic Heart Disease, OSA = Obstructive sleep apnea, COPD = Chronic obstructive pulmonary disease, T2DM = Type 2 Diabetes Mellitus, BMI = Body Mass Index, CKD = chronic kidney disease, CLD = Chronic Liver Disease | |

**Table 4: Comparative Characteristics for Baseline vs. Post-Baseline Liver Metastasis Along with Patient Characteristics for All Controls**

|  | **Proportion of LM Cases, N = 3333** | | | **Patient Characteristics of all Control** |
| --- | --- | --- | --- | --- |
| **Characteristic** | **Baseline LM 2,066** | **Post baseline LM**  **1,267** | **p-value***^1^* | **Control 9,622** |
| **Age (years), Median (Q1, Q3)** | — | — |  | 73.0 (65.0, 80.0) |
| **Age Group, n (%)** |  |  | <0.001 |  |
| *45-60* | 232(11.2%) | 218(17.2%) |  | 1,136 (12%) |
| *61-74* | 988(47.8%) | 652(51.5%) |  | 4,319 (45%) |
| *>= 75* | 846(40.9%) | 397(31.3%) |  | 4,167 (43%) |
| **Ethnicity, n (%)** |  |  | 0.7 |  |
| *Not Hispanic or Latino* | 1,581(76.5%) | 956(75.5%) |  | 7,779 (81%) |
| *Hispanic or Latino* | 25(1.2%) | 19(1.5%) |  | 278 (2.9%) |
| *Unknown* | 460(22.3%) | 292(23.0%) |  | 1,565 (16%) |
| **Race, n (%)** |  |  | 0.9 |  |
| *White* | 1,571(76.0%) | 952 (75.1%) |  | 7,208 (75%) |
| *Black or African American* | 108(5.2%) | 72(5.7%) |  | 734 (7.6%) |
| *Other Race* | 28(1.4%) | 20(1.6%) |  | 398 (4.1%) |
| *Unknown* | 359(17.4%) | 223(17.6%) |  | 1,282 (13%) |
| **Sex, n (%)** |  |  | 0.9 |  |
| *Female* | 1,011(48.9%) | 624(49.3%) |  | 5,011 (52%) |
| *Male* | 1,055(51.1%) | 643(50.7%) |  | 4,611 (48%) |
| **Region, n (%)** |  |  | 0.2 |  |
| *Midwest* | 74(3.6%) | 55(4.3%) |  | 745 (7.7%) |
| *Northeast* | 33(1.6%) | 21(1.7%) |  | 325 (3.4%) |
| *South* | 82(4.0%) | 61(4.8%) |  | 1,206 (13%) |
| *Unknown* | 1,790(86.6%) | 1,062(83.8%) |  | 6,676 (69%) |
| *West* | 87(4.2%) | 68(5.4%) |  | 670 (7.0%) |
| **Distant Metastases/Node, n (%)** | 489(23.7%) | 123(9.7%) | <0.001 | 670 (7.0%) |
| **Primary Cancer Location, n (%)** |  |  | <0.001 |  |
| *Body/Tail* | 313(15.2%) | 165(13.0%) |  | 1,192 (12%) |
| *Head* | 281(13.6%) | 344(27.2%) |  | 1,998 (21%) |
| *Other* | 71(3.4%) | 61(4.8%) |  | 524 (5.4%) |
| *Unspecified* | 1,401(67.8%) | 697(55.0%) |  | 5,908 (61%) |
| **BMI Group, n (%)** |  |  | 0.3 |  |
| *Normal* | 734(35.5%) | 456(36.0%) |  | 3,464 (36%) |
| *Underweight* | 76(3.7%) | 28(2.2%) |  | 490 (5.1%) |
| *Overweight* | 654(31.7%) | 393(31.0%) |  | 3,185 (33%) |
| *Obese (Class I)* | 374(18.1%) | 239(18.9%) |  | 1,510 (16%) |
| *Obese (Class II)* | 133(6.4%) | 91(7.2%) |  | 615 (6.4%) |
| *Obese (Class III)* | 95(4.6%) | 60(4.7%) |  | 358 (3.7%) |
| **BMI, Median (Q1, Q3)** | — | — |  | 26.1 (22.8, 30.2) |
| **CKD, n (%)** | 526(25.5%) | 219(17.3%) | <0.001 | 2,163 (22%) |
| **T2DM, n (%)** | 942(45.6%) | 541(42.7%) | 0.1 | 4,195 (44%) |
| **Hepatitis, n (%)** | 62(3.0%) | 33(2.6%) | 0.5 | 275 (2.9%) |
| **Hypertension, n (%)** | 1,665(80.6%) | 954(75.3%) | <0.001 | 7,478 (78%) |
| **CLD, n (%)** | 113(5.5%) | 42(3.3%) | 0.004 | 454 (4.7%) |
| **Hyperlipidemia, n (%)** | 1,558(75.4%) | 877(69.2%) | <0.001 | 6,801 (71%) |
| **OSA, n (%)** | 401(19.4%) | 249(19.7%) | 0.9 | 1,726 (18%) |
| **COPD, n (%)** | 400(19.4%) | 196(15.5%) | 0.004 | 1,739 (18%) |
| **Anxiety, n (%)** | 619(30.0%) | 343(27.1%) | 0.074 | 2,405 (25%) |
| **IHD, n (%)** | 612(29.6%) | 299(23.6%) | <0.001 | 2,616 (27%) |
| **Depression, n (%)** | 481(23.3%) | 262(20.7%) | 0.08 | 1,813 (19%) |
| **Obesity, n (%)** | 630(30.5%) | 371(29.3%) | 0.5 | 2,530 (26%) |
| **Cancer, n (%)** | 404(19.6%) | 236(18.6%) | 0.5 | 1,619 (17%) |
| **Abdominal Pain, n (%)** | 1,339(64.8%) | 808(63.8%) | 0.5 | 5,376 (56%) |
| **Dyspnea, n (%)** | 679(32.9%) | 331(26.1%) | <0.001 | 2,900 (30%) |
| **Anemia, n (%)** | 971(47.0%) | 436(34.4%) | <0.001 | 3,848 (40%) |
| *^1^*Pearson's Chi-squared test; Fisher's Exact Test for Count Data with simulated p-value (based on 2000 replicates); Fisher's exact test | | | | |
| Abbreviations: CI = Confidence Interval, HR = Hazard Ratio, IHD = Ischemic Heart Disease, OSA = Obstructive sleep apnea, COPD = Chronic obstructive pulmonary disease, T2DM = Type 2 Diabetes Mellitus | | | | |

**Table 5: Distribution of Lab Function Test for Liver Metastasis (Cases) and Subjects Free of Liver Metastases (Control)**

| Lab Test (unit) | Total Cases, N = 2920 | Total Control, N = 4938 | Cases Median  (IQR) | Control Median  (IQR) | p-value | Hodges Lehmann Estimate^1^ |
| --- | --- | --- | --- | --- | --- | --- |
| Direct Bilirubin (mg/dL) | 1,935 | 2231 | 0.7 (1.38) | 0.6 (0.6) | <0.0001 | 0.1 |
| Total Bilirubin (mg/dL) | 1,971 | 2262 | 0.8 (1.4) | 0.6 (0.7) | <0.0001 | 0.2 |
| Alkaline Phosphatase (ALP) (U/L) | 1,899 | 2140 | 163 (274) | 92 (91.25) | <0.0001 | 47 |
| Aspartate Aminotransferase (AST) (U/L) | 1,924 | 2191 | 35 (54) | 24 (27) | <0.0001 | 7 |
| Alanine Aminotransferase (ALT) (U/L) | 1,918 | 2183 | 34 (54) | 25 (36) | <0.0001 | 5 |
| Albumin (g/dL) | 1,972 | 2263 | 3.4 (1.1) | 3.8 (1) | <0.0001 | -0.3 |
| Total Protein (g/dL) | 1,909 | 2185 | 6.6 (1.2) | 6.8 (1) | <0.0001 | -0.2 |
| Serum Lipase (U/L) | 1,120 | 1001 | 53 (115.25) | 58 (124) | NS | -4 |
| Serum Pancreatic Amylase (U/L) | 196 | 215 | 58.5 (184) | 53 (118.5) | NS | 5 |
| Prothrombin Time (s) | 1,168 | 995 | 13.9 (3.3) | 13.1 (2.45) | <0.0001 | 1 |

**Figure S1: Frequency of Primary Pancreatic Cancer Location**

Frequency of Primary Tumor location

|  | Frequency |
| --- | --- |
| Body | 649 |
| Duct | 127 |
| Head | 1593 |
| Other Parts | 226 |
| Overlapping Sites | 90 |
| Tail | 499 |
| Unspecified | 4950 |

**Figure S2: Distribution of observed and imputed BMI values.**

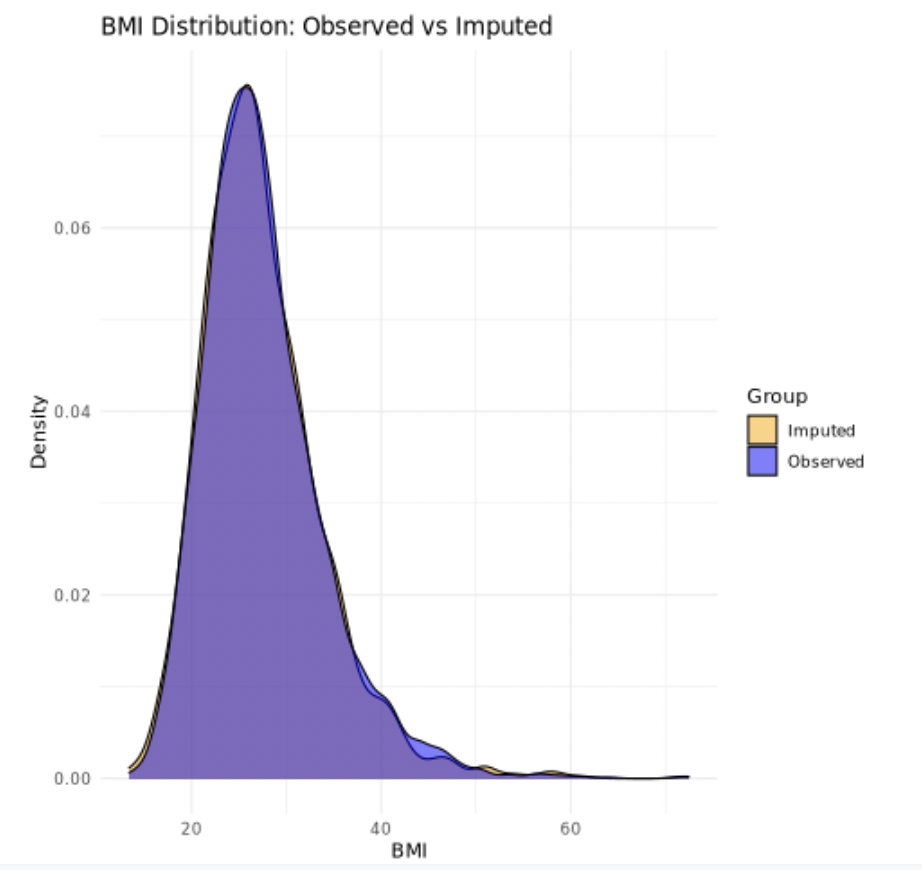

**Figure S3: Kaplan–Meier survival curves illustrating time to LM and the composite outcome (LM or death)**

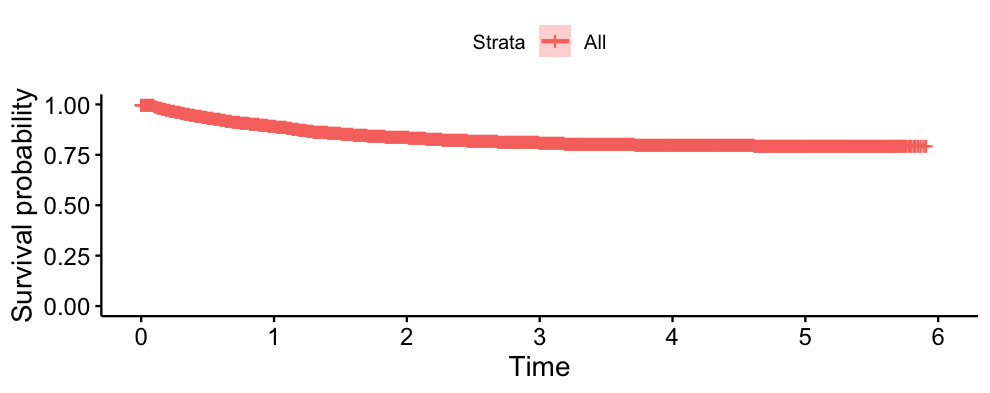

Figure 3.1. Kaplan–Meier Curve for Time to Liver Metastasis

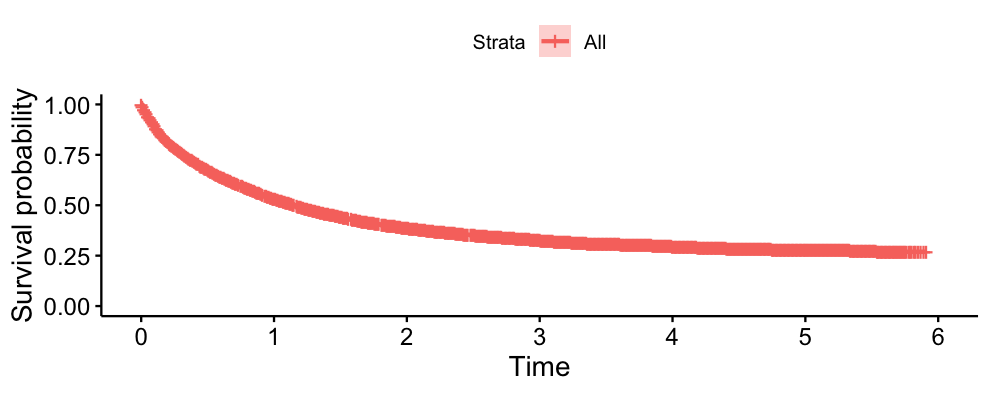

Figure 3.2. Kaplan–Meier Curve for Composite Outcome of Liver Metastasis or Death

eFigure3
